# Lactate Cut-offs for 28-day Mortality in Septic Shock

**DOI:** 10.64898/2026.02.08.26345840

**Authors:** Sophie-Theres Wanka, Alexander Hermann, Max Lenz, Christian Hengstenberg, Peter Schellongowski, Thomas Staudinger, Robert Zilberszac

**Affiliations:** Medical University of Vienna, Department of Medicine I, Vienna, Austria; Medical University of Vienna, Department of Cardiology, Vienna, Austria

**Keywords:** sepsis, septic shock, lactate, lactate cut-offs, 28-day mortality

## Abstract

**Background/Objectives:** Early lactate is widely used to risk-stratify septic shock, yet clinically actionable cut-offs for 28-day mortality remain uncertain.

**Methods:** In a single-centre study conducted across two intensive care units, we analysed 84 adults with septic shock identified within 24 hours of intensive care unit admission. The primary endpoint was 28-day mortality. Four lactate metrics obtained during the first 24 hours were evaluated: first (admission) lactate, last lactate, peak lactate, and lactate clearance from first to last. Associations were tested using logistic regression with and without adjustment for the Simplified Acute Physiology Score 3; discrimination was assessed by area under the receiver-operating characteristic curve (AUROC), and optimal cut-offs were defined by the Youden index.

**Results:** Thirty-nine of 84 patients (46.4%) died by day 28. Higher absolute lactate values were independently associated with death (adjusted odds ratio (OR) per 1 mmol/L increase: First 1.47, p < 0.001; Last 1.41, p = 0.002; Peak 1.39, p < 0.001), whereas lactate clearance was not (OR 0.65, p=0.202). Discrimination was moderate to good for peak (AUROC 0.817), first (0.791), and last (0.757) lactate, and poor for clearance (0.577). Youden-derived thresholds provided pragmatic trade-offs: First 3.55 mmol/L (sensitivity 0.821, specificity 0.689), Last 3.15 mmol/L (0.567, 0.864), and Peak 3.55 mmol/L (0.973, 0.556). Kaplan-Meier curves using these cut-offs showed early and sustained separation.

**Conclusions:** In adults with septic shock, absolute lactate measures showed higher observed discrimination for 28-day mortality than lactate clearance. The identified thresholds require external validation before clinical use.

## 1. Introduction

Sepsis affects millions of patients worldwide and remains a leading cause of mortality in critical care [1,2]. According to the Sepsis-3 definition, sepsis is characterized by life-threatening organ dysfunction resulting from a dysregulated host response to infection, while septic shock represents its most severe manifestation, associated with profound circulatory and metabolic abnormalities and high short-term mortality [3]. Disease severity and outcome in sepsis are influenced by both hyperinflammatory and immunosuppressive host responses, ultimately leading to tissue hypoperfusion and organ failure [4,5]. Early identification of high-risk patients is therefore essential. Among available clinical tools, severity scores such as the Simplified Acute Physiology Score 3 (SAPS 3) and biochemical markers have an important role in early risk stratification [6,7]. One sensitive marker is serum lactate, which reflects cellular metabolism and serves as a predictor of mortality [6,8]. Elevated lactate levels in sepsis and septic shock are associated with increased mortality and appear to reflect disease severity resulting from a combination of tissue hypoxia and stress-related metabolic responses [4]. According to the Sepsis-3 definition, a serum lactate concentration above 2 mmol/L is required for the diagnosis of septic shock [3].

However, measurement of serum lactate alone is not sufficient to establish the diagnosis of sepsis [6]. A systematic review of 28 studies concluded that serum lactate concentrations can serve as indicators of outcome, although no specific cut-off with acceptable sensitivity and specificity to predict hospital mortality or organ failure could be determined. Nonetheless, achieving normalization of lactate levels was recommended [9]. In our recent study, we demonstrated that the combination of SAPS 3 and early lactate measurements (first, peak, and last) provides markedly improved prognostic accuracy for 28-day mortality compared with SAPS 3 alone, with initial lactate showing the strongest predictive value (Area Under Curve ≈ 0.80) [10]. Building on these findings, the present study aims to determine candidate cut-off values for early lactate metrics in patients with septic shock, to enable more reliable early risk stratification and outcome prediction.

## 2. Materials and Methods

### 2.1. Study Design

We conducted a retrospective cohort study of patient data from two intensive care units situated at the Medical University of Vienna, Vienna General Hospital from January 2017 to December 2019. Consecutive ICU admissions were screened; of 998 unique ICU stays, 84 patients fulfilled the shock criteria within 24 hours and formed the study cohort.

### 2.2. Study Population

Patients included in this study were adults (≥18 years) admitted to one of the participating intensive care units during the study period who fulfilled the Sepsis-3 definition of septic shock. The diagnosis required hypotension requiring vasopressor support to maintain a mean arterial pressure of at least 65 mmHg and a serum lactate concentration above 2 mmol/L despite adequate fluid resuscitation. Exclusion criteria included incomplete records, pre-existing limitations of therapy, non-septic or mixed shock aetiology, and secondary transfers without complete admission data.

### 2.3. Lactate Parameters and Outcome

Lactate was analysed as four prespecified metrics derived from routine arterial/venous blood gas measurements obtained during the first 24 hours after ICU admission:

1. First = the first lactate value measured after ICU admission;
2. Last = the last lactate value measured within 24 hours;
3. Peak = the maximum lactate value within 24 hours;
4. Lactate clearance (%) between First and Last, defined as ((First − Last)/First) × 100; negative values denote a rise in lactate.

First, Last, and Peak lactate were expressed in millimoles per litre (mmol/L), whereas lactate clearance was expressed as a percentage. Lactate availability varied across metrics (First n=84, Peak n=82, Last/Clearance n=74). Analyses of Last and clearance used complete-case data without imputation; denominators are reported accordingly.

The sole outcome for this study was 28-day all-cause mortality, ascertained from hospital records and verified against ICU information systems.

Baseline variables included age, sex, Body Mass Index, comorbidities, and SAPS 3 at admission and were extracted from the ICU database and electronic records. SAPS 3 served as the a priori severity covariate in adjusted models.

### 2.4. Statistical Analysis

We fitted univariable and SAPS 3–adjusted logistic regression models with 28-day mortality as the dependent variable and lactate metrics as predictors, reporting odds ratios with 95% confidence intervals.

Discriminative performance for 28-day mortality was evaluated by area under the receiver-operating characteristic curve (AUROC) for each lactate metric and for SAPS 3. To identify candidate thresholds, we derived:

- the Youden-optimal cut-off (maximising sensitivity + specificity − 1), and
- a high-sensitivity cut-off constrained to specificity ≥ 50%.

For prespecified cut-points, we calculated sensitivity, specificity, positive predictive value (PPV), and negative predictive value (NPV) using observed cohort prevalence.

While the primary modelling framework was logistic regression for a fixed 28-day endpoint, we also generated Kaplan-Meier curves to visualise survival-curve separation for Youden-derived thresholds of first, last, and peak lactate and for an exploratory clearance threshold. Survival distributions were compared using the log-rank test.

All analyses were repeated after stratification by SAPS 3 > 80 vs. ≤ 80.

Analyses used nominal two-sided p-values (α = 0.05); missing exposure data were handled by complete-case analysis without imputation.

### 2.5. Artificial Intelligence Statement

AI tools (ChatGPT, OpenAI) were used solely to support language editing, improve clarity and provide technical guidance for SPSS procedures. No AI tools were used to generate, analyse, or interpret study data or results. All AI-assisted content was critically reviewed and verified by the authors, who take full responsibility for the final content of the manuscript.

### 2.6. Reporting Guidelines

This study was reported in accordance with the Strengthening the Reporting of Observational Studies in Epidemiology (STROBE) guidelines.

## 3. Results

### 3.1. Patient Characteristics

Baseline characteristics are displayed in Table 1. Patients included in our study (n = 84) had a median age of 62 (22-83) years and a balanced sex distribution (47.6% female). The median SAPS 3 on admission was 83.0 (49-122), with higher values observed among non-survivors (87.0 (59-122)) compared with survivors (80.0 (49-115)). The median ICU length of stay was 6 days (1–76), longer in survivors (13 days) than in non-survivors (2 days). Median Body Mass Index was 25.2 kg/m² (17-62). Immunosuppression (defined as having hematological malignancies, active solid tumor, solid-organ transplant, acquired immunodeficiency syndrome, or long-term or high-dose corticosteroid or immunosuppressant use [11]) and arterial hypertension were the most common comorbidities (each 40.5%), followed by malignancy (27.4%), chronic obstructive pulmonary disease (21.4%), diabetes mellitus (17.9%), chronic kidney disease (13.1%), coronary artery disease (10.7%), and peripheral arterial disease (8.3%).

**Table 1.**
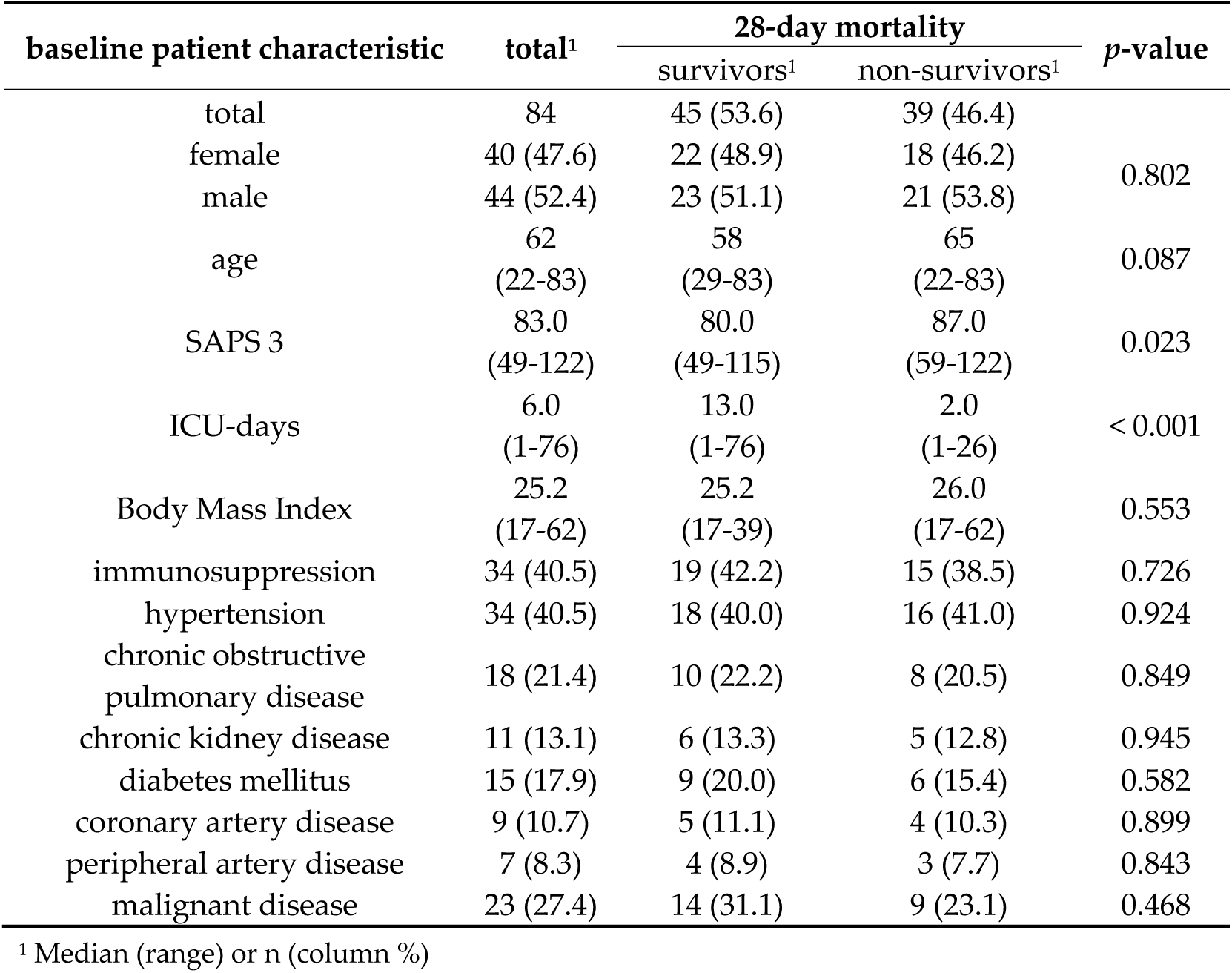
Baseline patient characteristics.

Among 84 patients who met criteria for septic shock within 24 h of ICU admission, 39 deaths occurred within 28 days (46.4%); the remaining 45 patients were censored alive at day 28. All analyses below exclusively address 28-day mortality. Baseline characteristics by 28-day outcome were broadly comparable, with a numerically higher acuity among non-survivors (median age 65 (22-83) vs. 58 (29-83) years; SAPS 3 87.0 (59-122) vs. 80.0 (49-115)).

Availability of lactate-based metrics varied slightly by variable: analyses using first lactate included all 84 patients, peak lactate within 24 hours included 82, and last lactate within 24 hours and lactate clearance (from First to Last) included 74 patients, reflecting missingness late in the first 24 hours. These denominators were used consistently across the corresponding Receiver Operating Characteristic, threshold, and time-to-event evaluations.

### 3.2. SAPS 3 and 28-day Outcome

The SAPS 3 was markedly higher among non-survivors. Among patients with a SAPS 3 below 60.0, one patient died while eight survived, corresponding to a survival rate of 88.9%. In the range of 60.0 to 69.0, 11 of 17 patients survived (64.7%). From a SAPS 3 of 80.0 and above, mortality increased substantially across all subgroups. At scores above 90.0, only 7 of 21 patients survived, corresponding to a survival rate of 33.3%.

### 3.3. Lactate Levels and 28-day Outcome

Early lactate exposure was higher among 28-day non-survivors across all absolute measures. Median values (survivors vs. non-survivors) were: First 2.7 (0.9-10.5) vs. 5.9 (0.9-22.0) mmol/L p < 0.001, Last 2.1 (0.9-9.6) vs. 4.1 (1.2-18.0) mmol/L p < 0.001, and Peak 3.2 (2.1-9.9) vs. 9.3 (2.7-27.0) mmol/L p < 0.001. Lactate clearance was higher among survivors (median 25.5% (−256.0 to 84.0%) vs. 15.0% (−275.0 to 91.0%) p = 0.266). Distributions were right-skewed with wide ranges (e.g., Peak overall 2.1–27.0 mmol/L) (Table 2).

**Table 2.** Lactate variables and 28-day Mortality.

| unit | total<sup>1</sup> | 28-day mortality |  | p-value |
| --- | --- | --- | --- | --- |
|  |  | survivors<sup>1</sup> | non-survivors<sup>1</sup> |  |
| First<br>(mmol/L) | 4.0<br>(0.9-22.0) | 2.7<br>(0.9-10.5) | 5.9<br>(0.9-22.0) | < 0.001 |
| Last<br>(mmol/L) | 2.3<br>(0.9-18.0) | 2.1<br>(0.9-9.6) | 4.1<br>(1.2-18.0) | < 0.001 |
| Peak<br>(mmol/L) | 4.5<br>(2.1-27.0) | 3.2<br>(2.1-9.9) | 9.3<br>(2.7-27.0) | < 0.001 |
| Lactate<br>clearance<sup>2</sup> | 22.5<br>(-275.0 to 91.0) | 25.5<br>(-256.0 to 84.0) | 15.0<br>(-275.0 to 91.0) | 0.266 |

<sup>1</sup> Median (range); <sup>2</sup> %

### 3.4. Logistic Regression Analysis

In univariable analyses (per 1 mmol/L increase), First, Last, and Peak lactate were each associated with higher odds of 28-day death: First OR 1.496 (95% Confidence Interval 1.196–1.871; p < 0.001), Last OR 1.428 (1.156–1.764; p < 0.001), and Peak OR 1.410 (1.183–1.681; p < 0.001). Lactate clearance showed the expected inverse directionality but was not statistically significant (OR 0.636 per 1.0-unit increase in lactate clearance, with clearance expressed as a proportion; 0.325–1.245; p = 0.187) (Table 3). For context, SAPS 3 alone was positively associated with 28-day mortality (OR per point 1.034, 1.004–1.066; p = 0.028).

**Table 3.** Univariable Logistic Regression Analysis of Lactate variables and 28-day Mortality.

|  | Regression Coefficient | Standard Error | p-value | Odds-Ratio | 95% Confidence Interval |  |
| --- | --- | --- | --- | --- | --- | --- |
| First | 0.402 | 0.114 | < 0.001 | 1.496 | 1.196 | 1.871 |
| Last | 0.356 | 0.108 | < 0.001 | 1.428 | 1.156 | 1.764 |
| Peak | 0.344 | 0.090 | < 0.001 | 1.410 | 1.183 | 1.681 |
| Lactate clearance<sup>1</sup> | -0.452 | 0.343 | 0.187 | 0.636 | 0.325 | 1.245 |

<sup>1</sup> Lactate clearance was analyzed as a proportion; the reported odds ratio corresponds to a 1.0-unit increase in the proportion.

Each absolute lactate metric retained an independent association with 28-day mortality after adjustment for SAPS 3: First OR 1.472 (p < 0.001), Last OR 1.413 (p = 0.002), and Peak OR 1.391 (p < 0.001). In these models, SAPS 3 itself was not significant (p = 0.405, 0.685, and 0.437, respectively). Lactate clearance remained non-significant after adjustment (OR 0.648 per 1.0-unit increase in the clearance proportion; p = 0.202) (Table 4).

**Table 4.** Multivariable Logistic Regression Analysis.

|  | Regression Coefficient | Standard Error | p-value | Odds-Ratio | 95% Confidence Interval |  |
| --- | --- | --- | --- | --- | --- | --- |
| SAPS 3 | 0.014 | 0.017 | 0.405 | 1.014 | 0.981 | 1.049 |
| First | 0.387 | 0.116 | < 0.001 | 1.472 | 1.174 | 1.847 |
| SAPS 3 | 0.008 | 0.019 | 0.685 | 1.008 | 0.971 | 1.045 |
| Last | 0.346 | 0.109 | 0.002 | 1.413 | 1.141 | 1.750 |
| SAPS 3 | 0.014 | 0.017 | 0.437 | 1.014 | 0.980 | 1.049 |
| Peak | 0.330 | 0.091 | < 0.001 | 1.391 | 1.164 | 1.661 |
| SAPS 3 | 0.023 | 0.017 | 0.164 | 1.024 | 0.991 | 1.058 |
| Lactate clearance<sup>1</sup> | -0.434 | 0.340 | 0.202 | 0.648 | 0.333 | 1.262 |

<sup>1</sup> Lactate clearance was analyzed as a proportion; the reported odds ratio corresponds to a 1.0-unit increase in the proportion.

### 3.5. Receiver Operating Characteristic Analysis

Receiver-operating characteristic analyses demonstrated moderate-to-good discrimination for absolute lactate measures and inferior performance for clearance. AUROC values were: Peak 0.817, First 0.791, Last 0.757; Lactate clearance 0.577 (p = 0.266) and SAPS 3 0.650. All three absolute lactate AUROCs were highly significant (p < 0.001) (Figure 1).

**Figure 1.**
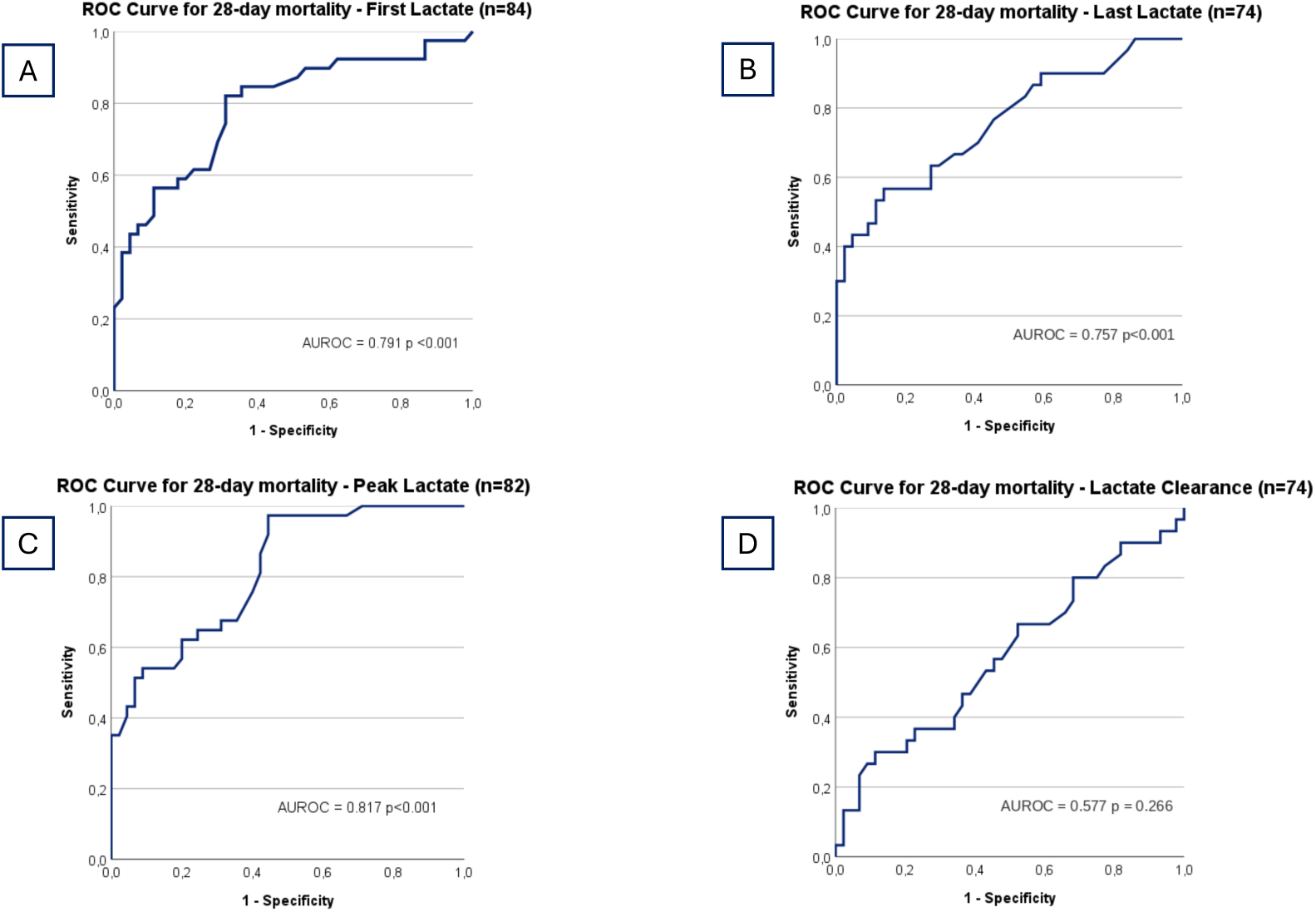
Receiver Operating Characteristic Curves of Lactate variables predicting 28-day Mortality; (A) First, (B) Last, (C) Peak, (D) Lactate clearance

### 3.6. Lactate Cut-offs

Youden-optimal and high-sensitivity cut-offs highlighted pragmatic risk-stratification thresholds (sens/spec):

- First: 3.55 mmol/L (Youden-optimal) 0.821/0.689; a higher-sensitivity alternative at 2.80 mmol/L achieved 0.846/0.556.
- Last: 3.15 mmol/L (Youden-optimal) 0.567/0.864; a higher-sensitivity but moderate specificity was found at 2.15 mmol/L with 0.767/0.545.
- Peak: 3.55 mmol/L (Youden-optimal) 0.973/0.556; the 3.35 mmol/L cut-off yielded 0.973/0.533.

Expanded operating characteristics (sensitivity, specificity, PPV, NPV) across clinically familiar thresholds corroborated these patterns. For example, Peak > 3.55 mmol/L achieved sensitivity 97.3% and NPV 96.2%, whereas Last > 3.15 mmol/L favored specificity (86.4%) with PPV ≈ 74% (Table 5 and Table 6).

**Table 5.** Cut-off values of Lactate variables for 28-day Mortality.

|  | Cut-off<sup>1</sup> | Sensitivity | Specificity | Youden Index |
| --- | --- | --- | --- | --- |
| First | > 2.80 | 0.846 | 0.556 | 0.402 |
| First | > 3.55 | 0.821 | 0.689 | 0.509 |
| Last | > 2.15 | 0.767 | 0.545 | 0.312 |
| Last | > 3.15 | 0.567 | 0.864 | 0.430 |
| Peak | > 3.35 | 0.973 | 0.533 | 0.506 |
| Peak | > 3.55 | 0.973 | 0.556 | 0.529 |
| Lactate clearance | < 25.5 | 0.600 | 0.500 | 0.100 |
| Lactate clearance | ≤ -46.5 | 0.300 | 0.886 | 0.186 |

<sup>1</sup> Cut-offs are expressed in mmol/L for First, Last, and Peak lactate and in % for lactate clearance

**Table 6.**
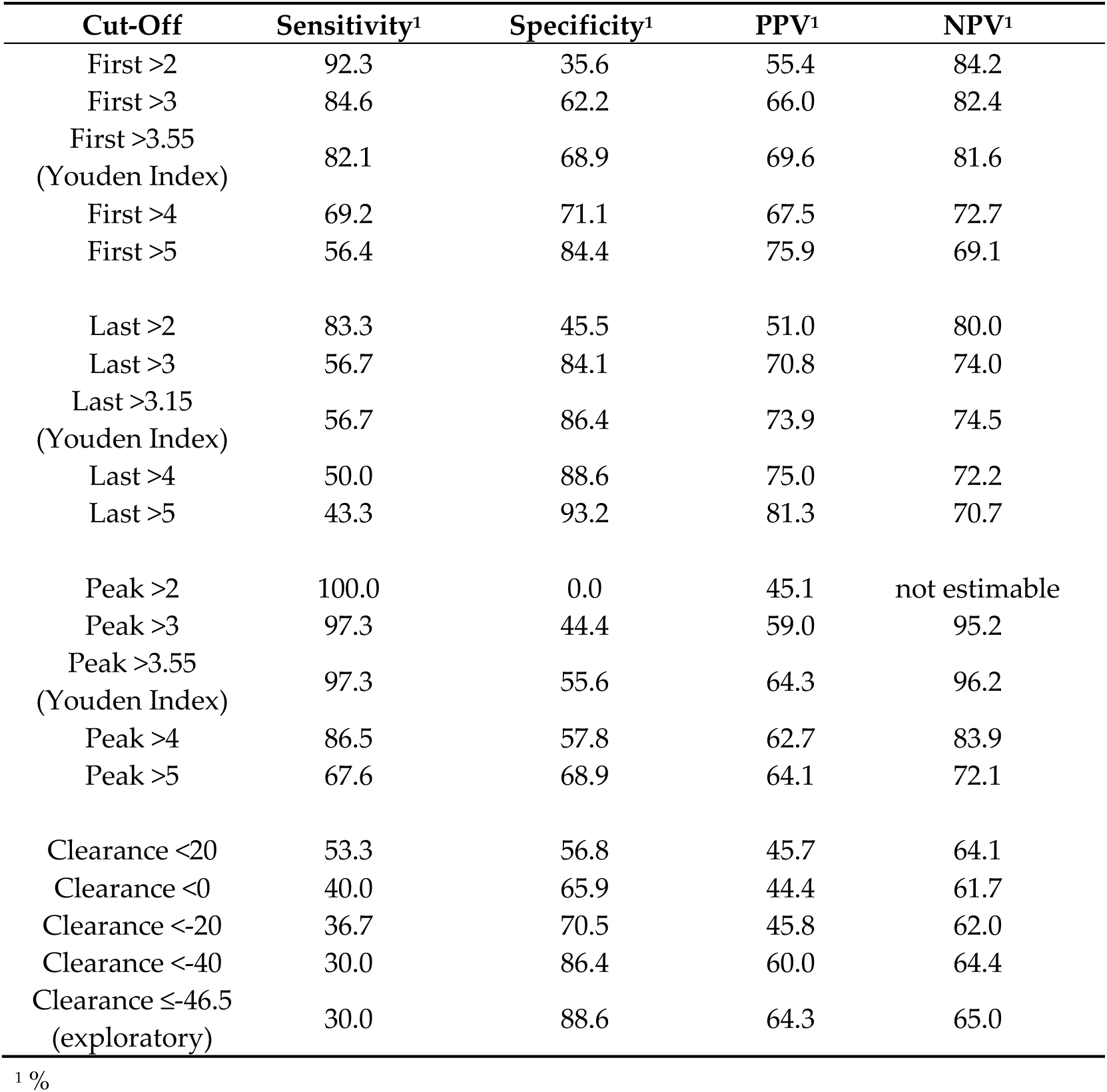
Predictive performance of Lactate Cut-offs for 28-day Mortality.

| Cut-Off | Sensitivity<sup>1</sup> | Specificity<sup>1</sup> | PPV<sup>1</sup> | NPV<sup>1</sup> |
| --- | --- | --- | --- | --- |
| First >2 | 92.3 | 35.6 | 55.4 | 84.2 |
| First >3 | 84.6 | 62.2 | 66.0 | 82.4 |
| First >3.55<br>(Youden Index) | 82.1 | 68.9 | 69.6 | 81.6 |
| First >4 | 69.2 | 71.1 | 67.5 | 72.7 |
| First >5 | 56.4 | 84.4 | 75.9 | 69.1 |
| Last >2 | 83.3 | 45.5 | 51.0 | 80.0 |
| Last >3 | 56.7 | 84.1 | 70.8 | 74.0 |
| Last >3.15<br>(Youden Index) | 56.7 | 86.4 | 73.9 | 74.5 |
| Last >4 | 50.0 | 88.6 | 75.0 | 72.2 |
| Last >5 | 43.3 | 93.2 | 81.3 | 70.7 |
| Peak >2 | 100.0 | 0.0 | 45.1 | not estimable |
| Peak >3 | 97.3 | 44.4 | 59.0 | 95.2 |
| Peak >3.55<br>(Youden Index) | 97.3 | 55.6 | 64.3 | 96.2 |
| Peak >4 | 86.5 | 57.8 | 62.7 | 83.9 |
| Peak >5 | 67.6 | 68.9 | 64.1 | 72.1 |
| Clearance <20 | 53.3 | 56.8 | 45.7 | 64.1 |
| Clearance <0 | 40.0 | 65.9 | 44.4 | 61.7 |
| Clearance <-20 | 36.7 | 70.5 | 45.8 | 62.0 |
| Clearance <-40 | 30.0 | 86.4 | 60.0 | 64.4 |
| Clearance ≤-46.5<br>(exploratory) | 30.0 | 88.6 | 64.3 | 65.0 |

<sup>1</sup> %

### 3.7. Kaplan-Meier Survival Analysis

Kaplan-Meier analyses using Youden-derived thresholds for First, Last and Peak lactate and an exploratory threshold for lactate clearance showed early and sustained separation of survival curves (Figure 2):

- First > 3.55 mmol/L: 32/46 deaths vs. 7/38 when ≤ 3.55 mmol/L (N = 84) p < 0.001.
- Last > 3.15 mmol/L: 17/23 deaths vs. 13/51 when ≤ 3.15 mmol/L (available-case N = 74) p < 0.001.
- Peak > 3.55 mmol/L: 36/56 deaths vs. 1/26 when ≤ 3.55 mmol/L (available-case N = 82) p < 0.001.
- Lactate clearance ≤ −46.5%: 9/14 deaths vs. 21/60 deaths among patients with clearance > −46.5% (available-case N = 74) log-rank p = 0.015.

**Figure 2.**
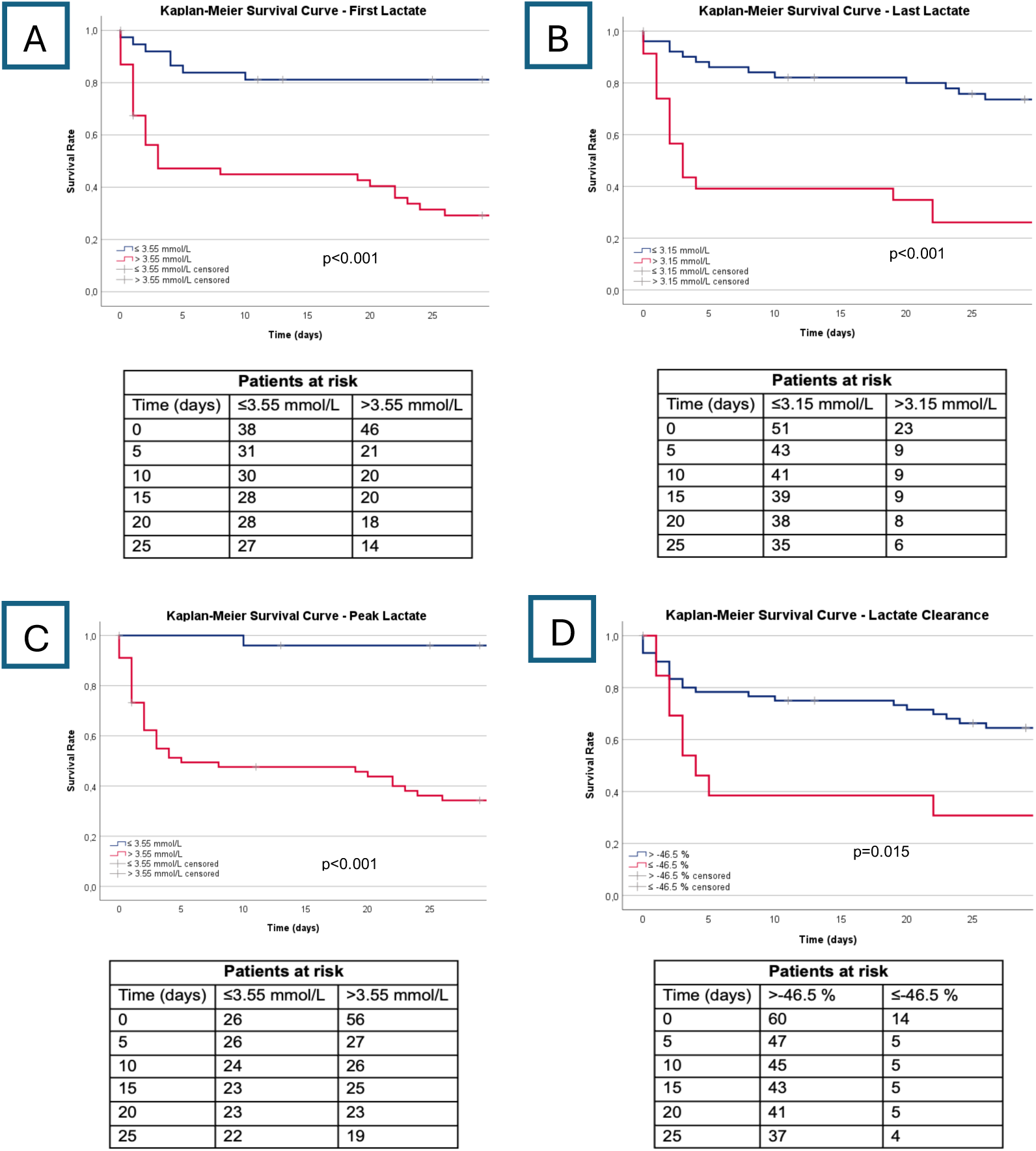
Kaplan-Meier Survival Curves for 28-day Mortality; (A) First, (B) Last, (C) Peak, (D) Lactate clearance

### 3.8. SAPS 3-Stratified Analysis

Diagnostic performance was broadly preserved across acuity strata (SAPS 3 > 80 vs. ≤ 80):

Cross-tabulation of Youden cut-offs. For First > 3.55 mmol/L, sensitivity/specificity were 84.6%/69.6% (SAPS 3 > 80) and 76.9%/68.2% (≤ 80). For Peak > 3.55 mmol/L, sensitivity remained near perfect with moderate specificity: 96.0%/52.2% (> 80) and 100.0%/59.1% (≤ 80). Last > 3.15 mmol/L emphasised specificity in both strata (81.8% and 90.9%) at modest sensitivity (55.6% and 58.3%).

Stratified AUROCs. AUROCs were like the overall analysis: First 0.806 (SAPS 3 > 80) and 0.760 (≤ 80); Last 0.698 and 0.822; Peak 0.814 and 0.807; clearance non-informative (0.449 and 0.364).

## 4. Discussion

We recently demonstrated that lactate measurements outperform SAPS 3 alone in predicting mortality in septic shock, with the exception of lactate clearance, which failed to show prognostic utility—likely because survivors tended to have low initial lactate and thus minimal potential for clearance, whereas patients with very high lactate frequently died early [10].

Building on these observations, the present study aimed to establish candidate cut-off values for different lactate variables to support early risk stratification in septic shock.

Across all analyses, absolute lactate values measured within the first 24 hours after ICU admission were strongly and independently associated with 28-day mortality, and this pattern was similar in patients with higher and lower SAPS 3 scores (> 80 vs. ≤ 80), with broadly similar observed discrimination in the two SAPS 3 strata. First and peak lactate values demonstrated the strongest discriminative ability, with clinically practical cut-offs around 3.3–3.6 mmol/L, while last lactate showed a slightly lower threshold (∼3.15 mmol/L). In contrast, lactate clearance demonstrated weak discrimination and no association with mortality after adjustment for SAPS 3. The similar cut-offs for first and peak lactate should be interpreted cautiously because peak lactate includes the first lactate measurement by definition.

In multivariable models, all absolute lactate measures remained associated with 28-day mortality after adjustment for SAPS 3. SAPS 3 was not statistically significant in these two-variable models.

These findings are consistent with current guideline recommendations. The Surviving Sepsis Campaign endorses lactate measurement as a key component of initial sepsis evaluation due to its strong association with mortality [6]. Previous studies have demonstrated that initial, peak, and last lactate values outperform traditional severity indices and that adding lactate into SAPS 3 improves risk prediction, particularly in septic shock [10,12,13]. Mortality increases non-linearly above critical lactate thresholds, supporting the clinical relevance of threshold-based risk stratification. However, guideline recommendations remain weak due to limited evidence and variability in lactate availability, and lactate alone is insufficient for diagnosis given limited sensitivity and specificity [6,13].

It is noteworthy that the 28-day mortality in this cohort (46%) was high compared with recent large-scale data, whereas control arm mortality often approximates 31.6% [14,15]. This likely reflects higher baseline illness severity, as indicated by markedly elevated SAPS 3 scores compared with typical international cohorts [16–18].

Furthermore, approximately 40% of patients were immunosuppressed, further distinguishing this population from major randomized trials such as ProCESS, ARISE, ProMISe, ADRENAL, and APROCCHSS, which enrolled patients with lower predicted mortality [14,15,19–23]. Thus, our findings emphasise the importance of considering real-world case-mix when interpreting sepsis trial outcomes and support lactate as a crucial prognostic marker in high-risk patient populations.

From a clinical perspective, our findings reinforce the central role of early lactate measurement in septic shock. The identified cut-offs, particularly the overlapping first and peak lactate thresholds near 3.5 mmol/L, are simple and rapidly obtainable. These thresholds may support exploratory risk stratification, but they require external validation before clinical implementation.

## 5. Study Limitations

This study has several limitations. First, it is a retrospective analysis from a single tertiary referral centre, and our cohort probably over-represents patients with high illness severity, including many immunosuppressed and oncological patients, which may limit generalisability. Second, missing data (particularly for late 24-hour lactate measurements) reduced sample size for some analyses. Third, the modest overall sample size increases the risk of statistical imprecision and may have limited our ability to perform more granular subgroup analyses. Finally, the retrospective design inherently carries susceptibility to unmeasured confounding and documentation variability. Despite these limitations, detailed review of clinical records and expert adjudication of doubtful cases should have improved the accuracy of septic shock classification and strengthened the observed associations.

## 6. Conclusions

Early absolute lactate levels within 24 hours—particularly peak and first values— were associated with 28-day mortality after adjustment for SAPS 3. The identified cut-offs separated survival curves in this cohort but require external validation. Lactate clearance adds little prognostic value for this endpoint, however serial assessment of lactate alongside the evolving clinical picture remains essential for detecting early deterioration and enhancing prognostic judgment. These findings support external validation of absolute lactate thresholds for risk stratification in larger multicentre cohorts.

## Author Contributions

Conceptualization, R.Z., A.H. and S.-T.W.; methodology, R.Z., A.H. and S.-T.W.; formal analysis, S.-T.W. and R.Z.; investigation, S.-T.W.; data curation, S.-T.W.; visualization, S.-T.W.; writing—original draft preparation, R.Z., A.H. and S.-T.W.; writing—review and editing, M.L., C.H., P.S., T.S. and R.Z.; supervision, R.Z. All authors have read and agreed to the published version of the manuscript.

## Institutional Review Board Statement

The study was conducted in accordance with the Declaration of Helsinki and approved by the Ethics Committee of the Medical University of Vienna (protocol code EK 1853/2019).

## Informed Consent Statement

Patient consent was waived due to the retrospective observational design of the study.

## Data Availability Statement

The data supporting the findings of this study are not publicly available due to their sensitive nature but are available from the corresponding author upon reasonable request.

## Acknowledgments

We thank Arthur Stoiber, Dominik Christ, Felix Meixner, Florian Chiari, Gregor Klemm, Leon Kiss and Sebastian Markart for their invaluable work in making this project happen.

## Abbreviations

The following abbreviations are used in this manuscript:

AUROC: Area Under the Receiver Operating Characteristic Curve
FIRST: First lactate
ICU: Intensive Care Unit
LAST: Last lactate
NPV: Negative Predictive Value
OR: Odds Ratio
PEAK: Peak lactate
PPV: Positive Predictive Value
SAPS 3: Simplified Acute Physiology Score 3

